# Healthcare Providers’ Acceptability of Breast Milk Donation and Banking, and Factors Influencing Adoption in Rwanda: A Mixed-Methods Study

**DOI:** 10.64898/2026.09.10.26362791

**Authors:** Marie Immaculee Dusingize, Peace Ingabire, Peace Kakibibi, Augustine Ndaimani, Olayinka Ibrahim, Rex Wong

## Abstract

Breast milk is essential for optimal neonatal nutrition and survival, particularly among preterm and low-birth-weight infants. When a mother’s own milk is unavailable, donor breast milk is the recommended alternative. However, Rwanda currently lacks accredited breast milk donation systems and milk banks, and little is known about healthcare providers’ perspectives on their potential implementation.

The main objective of the study was to assess the acceptability of breast milk donation and explore the potential factors that could influence its adoption among healthcare providers in two Rwandan district hospitals.

A convergent parallel design with a qualitative-dominant approach was conducted in the neonatology units of Kirehe District Hospital and Ruhengeri Level II Teaching Hospital, Rwanda. Quantitative data were collected from 41 healthcare providers using structured questionnaires, including a six-item Likert scale (0 = strongly disagree to 4 = strongly agree) to measure acceptability. Scores ≥ 2.4 were interpreted as indicating high acceptability. Qualitative data were obtained through in-depth interviews with 10 healthcare providers and analyzed using inductive thematic analysis. Quantitative data were analyzed using descriptive statistics and non-parametric tests.

The overall acceptability of breast milk donation was high, with 82.9% of healthcare providers expressing agreement (mean score: 2.83 ± 1.10). Most participants (97.6%) perceived donor breast milk as beneficial for neonatal health. Key barriers included cultural resistance, limited maternal awareness, and inadequate infrastructure, while facilitators were staff training, institutional policies, and community engagement. Qualitative findings highlighted that providers were generally familiar with the concept of breast milk donation but noted the absence of formal programs in Rwanda. They emphasized that cultural concerns often stem from limited knowledge rather than entrenched opposition and strongly endorsed the benefits for infant health and maternal wellbeing. Providers stressed that successful implementation would depend on awareness-raising, trust in safety procedures, and institutional support through training and clear guidelines.

Breast milk donation is highly acceptable among healthcare providers in Rwanda, with a mean score of 2.83 ± 1.10 on the 0–4 Likert scale. Providers strongly recognized its benefits for vulnerable infants. Addressing gaps in awareness, infrastructure, and policy frameworks, alongside culturally sensitive community education, will be critical for successful implementation. These findings support the feasibility of introducing breast milk banking in Rwanda and provide evidence to guide policy and program development.

## INTRODUCTION

The World Health Organization (WHO) recommends that all infants be exclusively breastfed for the first six months of life, with early initiation of breastfeeding within one hour of birth, followed by the timely introduction of nutritionally adequate complementary foods while continuing breastfeeding up to two years [1], [2]. However, globally, only about 44% of infants aged 0–6 months are exclusively breastfed between 2015 and 2020 [1], highlighting the persistent gap between recommendations and practice. This low implementation rate is influenced by multiple factors, including maternal employment, limited access to lactation support, cultural norms, early introduction of formulas, and healthcare system barriers that hinder optimal infant feeding practices [3], [4], [5], [6], [7], [8].

Alongside breastfeeding promotion efforts, donor breast milk and breast milk banking have expanded in recent years and have been associated with increased access to breast milk (BM) to hospitalized and preterm infants, as well as reductions in feeding interruptions and morbidity such as necrotizing enterocolitis (NEC) and infections [9], [10]. According to the WHO and UNICEF [1], [11], when a mother’s own milk cannot be provided, pasteurized donor breast milk from a breast milk bank is the most preferred alternative for vulnerable infants, provided that a safe, quality assured system is in place. Evidence from systematic reviews and country-level reports further suggests that milk banking, when integrated with lactation support, can increase the proportion of infants receiving breast milk and improve short-term neonatal outcomes [10], [12], [13]. Furthermore, emerging evidence also points to cost-effectiveness. For instance, donor milk supplementation has been associated with lower hospital and feeding costs compared to formula feeding in very-low-birth-weight infants, largely due to shorter hospital stays and fewer complications [14], [15].

Despite global progress, disparities persist. Across sub-Saharan Africa, breastfeeding outcomes remain suboptimal despite longstanding promotional efforts [16], [17]. With the highest infant mortality rates globally, the continent only has eight operational milk banks as of 2025, with facilities limited to South Africa, Kenya, Nigeria, Angola, Cameroon, Mozambique, Uganda, and Cape Verde [18]. While many countries have made progress in promoting early initiation of exclusive breastfeeding; social norms, myths, awareness, and cultural beliefs continue to influence how breast milk donation is perceived [19]. These underscore the importance of context sensitive approaches in promoting donor breast milk and milk banking across the region.

Furthermore, evidence from rural Rwanda indicates that nearly half (46.5%) of preterm and low- birth-weight infants face feeding difficulties, often accompanied by growth and developmental delays [20]. While feeding difficulties do not necessarily imply complete absence of breast milk, evidence from neonatal care settings shows that preterm infants are significantly less likely to receive their mother’s own milk. For example, a recent study among preterm infants with feeding challenges found that only 35.4% were receiving mother’s own milk at discharge, and very few were exclusively breastfed [21]. These lower rates reflect difficulties with breastfeeding initiation and sustained milk expression, as well as prolonged mother–infant separation, which together limit timely access to mother’s own milk among pre-term infants.

Despite the national commitments to improve neonatal outcomes, no breast milk banks currently exist, and the practice of breast milk donation remains largely unfamiliar. This gap poses challenges for neonatal units that often struggle to provide adequate feeding options when mothers are unable to breastfeed. Donor human milk and breast milk banking could help ensure that these vulnerable infants receive the protective benefits of human milk during the critical early period when maternal milk is unavailable or insufficient. Rwanda faces a substantial gap in feeding support for preterm infants, which could be addressed by establishing human milk banks, as implemented in other countries.

Health professionals are well-positioned to advocate for the use of donor human milk in this context. However, the acceptability of donor human milk among these professionals remains unknown, and the perceived barriers to and facilitators of human milk banking in Rwanda have not been previously documented. Understanding health professionals’ perceptions is essential to inform future policy and guide the design of effective implementation programs. This study thus aims to explore the acceptability of breast milk donation for neonatal feeding in two Rwandan hospitals, focusing on the perspectives, beliefs, and factors influencing this acceptability among healthcare providers. By integrating quantitative and qualitative approaches, this study provides novel insights into the feasibility of introducing donor milk and milk banking in Rwandan hospitals.

## METHODS

### Study Settings

This study was conducted in the neonatology departments of two district hospitals; Kirehe District Hospitals (KDH) and Ruhengeri Level Two Teaching Hospital (RL2TH). Both facilities serve rural conservative populations in Rwanda’s Eastern and Northern provinces, respectively. The hospitals were purposively selected because they represent typical resource-constrained district-level healthcare settings in Rwanda with relatively high neonatal admission volumes and established neonatal care services.

At the time of the study, KDH had approximately 22 total staff, while RL2TH had around 25 staff members who were working in neonatology. These included nurses, doctors, cleaners, expert mothers, customer care agents, and social service providers (Source: preliminary desk research in both hospitals).

### Design

A concurrent qualitative-dominant mixed-methods design was employed, in which both qualitative and quantitative data were collected simultaneously. The qualitative component explored healthcare providers’ perspectives on breast milk donation, while the quantitative component assessed the level of acceptability and the determinants influencing its potential adoption. The qualitative strand followed a descriptive qualitative design, aimed at generating a straightforward, data-near account of healthcare providers’ perspectives rather than testing a specific theoretical framework. This mixed approach enables triangulation of findings to provide a comprehensive understanding of the factors shaping the feasibility of breast milk donation and banking in Rwandan hospital settings.

### Population

The study targeted Healthcare providers working within the same and collaborating units, including doctors, nurses, midwives, expert mothers, and social workers.

### Sample

#### Inclusion criteria

1. Doctors, nurses, midwives, and social workers working in the neonatology departments of the two hospitals.
2. Expert Mothers at KDH: Experienced mothers trained under the Partners In Health/ Inshuti Mu Buzima (PIH/IMB) program to provide peer-support in neonatal care. Their role includes breastfeeding counselling, Kangaroo Mother Care guidance, and psychosocial support for families of preterm and low-birth-weight infants [22].

#### Exclusion criteria

Administrative personnel and staff not directly involved in the provision or support of neonatal care services

### Sample size

#### Quantitative sampling

At the time of the study, the neonatology staff comprised of 25 personnel at RL2TH and 22 at KDH. Given the limited staff strength and therefore small population size, the study employed a census sampling approach, aiming for 100% participation from all healthcare providers employed in the two neonatology departments. However, due to staff availability at the time of data collection, the research team was able to reach only 41 participants (87.2%).

#### Qualitative sampling

The qualitative sample consisted of 10 healthcare providers (including doctors, nurses, midwives, and expert mothers) from both hospitals, selected through a purposive sampling approach to ensure representation of key cadres directly involved in neonatal care. Participants were interviewed until data saturation was reached, the point at which no new information emerged from responses. Our findings indicated that near code saturation was reached around the eighth HCP interview, but further interviews were conducted to attain an acceptable level of analytical saturation, ensuring adequate depth and variation across participant group. This aligns with recent studies emphasizing that saturation should be linked to analytical adequacy [23]. Similar studies have also shown that a slightly larger number of interviews may be required to achieve meaningful saturation, particularly when participants represent diverse backgrounds and contexts [23], [24]. All 10 healthcare providers approached for the qualitative interviews agreed to participate; none declined or withdrew before completing their interview.

### Data collection

#### Data collection procedure

Before data collection, the instruments were pretested to assess clarity, flow, and the performance of data collectors. Findings from the pretest indicated that, while the tools were generally understandable, data collectors required additional training, particularly on conducting semi-structured interviews. A one-day refresher training was conducted to strengthen their skills. Beyond the one-day refresher training on semi-structured interviewing described above, data collectors had prior experience conducting in-depth interviews as part of quality improvement projects at the hospitals where they previously practiced as nurses.

Data collection occurred from 10^th^ June through 13^th^ August 2025. To recruit participant HCPs, data collectors visited the neonatal units where all eligible staff were physically present while doing their daily responsibility. Each HCP was approached individually. The data collectors first introduced themselves, provided a brief introduction and summary explanation of the study’s purpose, followed by a simple question to assess initial interest in participation. For HCPs who expressed willingness to contribute, the data collectors then proceeded to provide a full explanation of the study, including its objectives. They proceeded by describing the voluntary nature of participation, and reviewed all the components of the consent form, including the option to decline participation without consequences. For participants involved in the qualitative component, consent for audio-recording was included. Written notes were taken only for interviews where audio recording was declined; no supplementary field notes were kept for recorded interviews.

#### Quantitative data collection

Quantitative data were collected using structured questionnaires administered through Google Forms. All available participants; 41 HCPs including doctors, nurses, midwives, expert mothers, and direct social workers working in the neonatal units, completed the survey. Items focused on HCPs’ perspectives regarding the role, feasibility, integration of breast milk donation in neonatal care, and institutional related factors affecting acceptability. In addition, a Likert scale item assessed their overall acceptability, reported on a scale of 0-4.

#### Qualitative data collection

The qualitative component involved semi structured in-dept interviews with conveniently selected HCPs to gather detailed insights into their views on breast milk donation and its potential implementation in neonatal care. All interviews were conducted in Kinyarwanda, audio-recorded with the informed consent, and transcribed verbatim before translation into English for analysis. Interviews lasted between nine and 40 minutes (with an approximate mean of 22 minutes). Transcripts were not returned to participants for review or correction; this is noted as a limitation below.

Interviews were conducted by nurses, one male and one female, who were trained in qualitative research, held a Bachelor of Science in Nursing, were students in a Master of Public Health program at the time of the study, and were recruited as data collectors. Interviewers had no prior personal or professional relationship with the participating healthcare providers before the study. During recruitment, participants were informed of the study’s purpose and objectives, but interviewers did not disclose personal views on breast milk donation prior to the interviews.

Each participant was interviewed once; no repeat interviews were conducted, and interviews were conducted in a private space within each hospital’s neonatology unit (e.g., an unoccupied office or consultation room), away from patient care areas.

### Data collection tools

The data collection tools for both the quantitative and qualitative components were developed by adapting and tailoring instruments from similar studies conducted in African settings with economic and sociocultural contexts comparable to Rwanda, including Uganda, Kenya, Ghana, and South Africa [25], [26], [27], [28]. These tools were further refined to reflect the specific characteristics and cultural nuances of the Rwandan population and needs in neonatal care context.

Prior to full scale data collection, a pilot study was conducted to assess the clarity, sense, flow, and cultural appropriateness of the instruments. The pilot included five quantitative surveys with HCPs, 2 qualitative interviews. Feedback from the pilot indicated that tools were generally understandable; however, minor adjustments were necessary.

#### Quantitative data collection tool

The structured tool for healthcare providers focused on their professional perspectives on introducing breast milk donation in neonatal care settings (*S1 Appendix).* The first part covered demographics (job role, years of experience, and, if applicable, prior exposure to breast milk donation programs). The second part assessed understanding of milk banking, perceived benefits, and potential impact on neonatal outcomes. The third part explored opinions on establishing a milk bank, policy barriers, and institutional practicalities of implementation. The fourth part reviewed views on existing hospital policies and required institutional support. The fifth part comprised a six-item Likert acceptability scale [29]. This scale demonstrated high internal consistency (Cronbach’s α = 0.935) and assessed key domains of acceptability, including attitudes toward donor milk, perceived benefits, perceived safety, willingness to donate or use donor milk, and overall comfort with the intervention, thereby supporting both the reliability and validity of the measure.

### S1 Appendix. Structured questionnaire for Healthcare Providers

#### Qualitative data collection tool

The interview guide *(S2 Appendix)* for HCPs was designed to capture detailed professional insights regarding the feasibility and implementation of breast milk donation programs. The guide explored: awareness, attitudes towards breast milk donation, perceptions of cultural acceptance among mothers and the broader community, anticipated operational, logistical, and ethical challenges, and requirements for successful implementation in the hospital settings. The open-ended format allowed HCPs to elaborate on their practical experiences, perceived institutional readiness, and recommendations for program integration.

### S2 Appendix. Semi-structured interview guide for Healthcare Providers

Key Measures

### Dependent Variables

#### 1. Acceptability

Acceptability measured the extent to which participants positively received the concept of breast milk donation and breast milk banking. This was measured using a six-item Likert scale (0–4), Classified into High acceptability: ≥ 2.4 and Low acceptability: < 2.4

### Independent Variables

#### 1. Awareness

This variable assessed participants’ awareness of breast milk donation and breast milk banking. It captured whether individuals had ever been exposed to the concept which scored 1 or were entirely unfamiliar with it which scored 0, emphasizing the role of informational exposure in shaping attitudes and behaviors.

#### 2. Cultural Beliefs and Attitudes

These variables integrated cultural values, and individual perceptions that influenced participants’ interpretations of breast milk donation and banking. It reflected how culturally informed beliefs and personal attitudes affected perceptions of safety, appropriateness, and moral acceptability, ultimately shaping acceptance and decision-making regarding donation.

#### 3. Perceived Benefits

Perceived benefits captured the advantages of participants associated with breast milk donation, including infant health promotion and support for vulnerable babies. Understanding these perceived benefits helped explain motivational factors that encouraged acceptance and potential participation in milk donation programs.

#### 4. Perceived Barriers

Perceived barriers encompass the challenges or concerns that may limit acceptance or willingness to donate. These included fears about safety, misinformation, cultural restrictions, logistical difficulties, or lack of trust in milk banking systems, highlighting obstacles that interventions must address to optimize uptake.

## Data analysis

All quantitative analyses were conducted using IBM SPSS Statistics version 26. Descriptive and inferential statistics were applied in alignment with study objectives and approaches commonly used in maternal health and donor-milk acceptability research.

Healthcare providers’ years of neonatal-care experience were dichotomized (<3 vs. ≥3 years), reflecting evidence that three years of continuous exposure marks acquisition of foundational competence [12], [30]. Awareness and perception variables (e.g., exposure to breast milk donation, cultural resistance, support for program introduction, policy development) were coded categorically (“Yes”, “No”, “Unsure”) and summarized using frequencies and percentages.

Acceptability was measured with a six-item Likert scale (0 = strongly disagree to 4 = strongly agree; maximum score = 24). Reliability was assessed with Cronbach’s α (>0.70 acceptable), and the scale demonstrated high internal consistency. Individual scores were averaged, and the 0–4 range was interpreted in equal intervals: 0–0.8 = strongly disagree, 0.8–1.6 = disagree, 1.6– 2.4 = neutral, 2.4–3.2 = agree, 3.2–4 = strongly agree. For summary analyses, scores ≥2.4 were classified as “agree,” following cutoffs adapted from similar studies in East and Southern Africa [31], [32], [33]. This approach facilitated consistent interpretation across studies.

Inferential analyses used non-parametric tests due to violation of normality (Shapiro–Wilk). Mann–Whitney U tests compared median acceptability across two-category variables (e.g., hospital, years of experience), while Kruskal–Wallis H tests examined variables with more than two categories (e.g., job title, education, socioeconomic status). Pearson’s chi-square tests assessed associations between dichotomized acceptability (≥2.4 vs. <2.4) and perception indicators, as well as between sociodemographic variables and awareness/perception measures. Statistical significance was set at p < 0.05, and missing data (<5%) were handled by listwise deletion.

Qualitative data were analyzed using manual inductive thematic analysis, following Braun and Clarke’s six phases [34]. Transcripts were read for familiarization, openly coded, and independently reviewed by the two principal investigators, who iteratively developed and finalized a codebook. Codes were condensed into categories and themes through consensus. The final coding structure, including themes, subthemes, categories, and codes, is presented in Figure 2 (indexed in the results section). Coding and thematic analysis were conducted manually, without the use of qualitative data analysis software.

**Fig 1.**
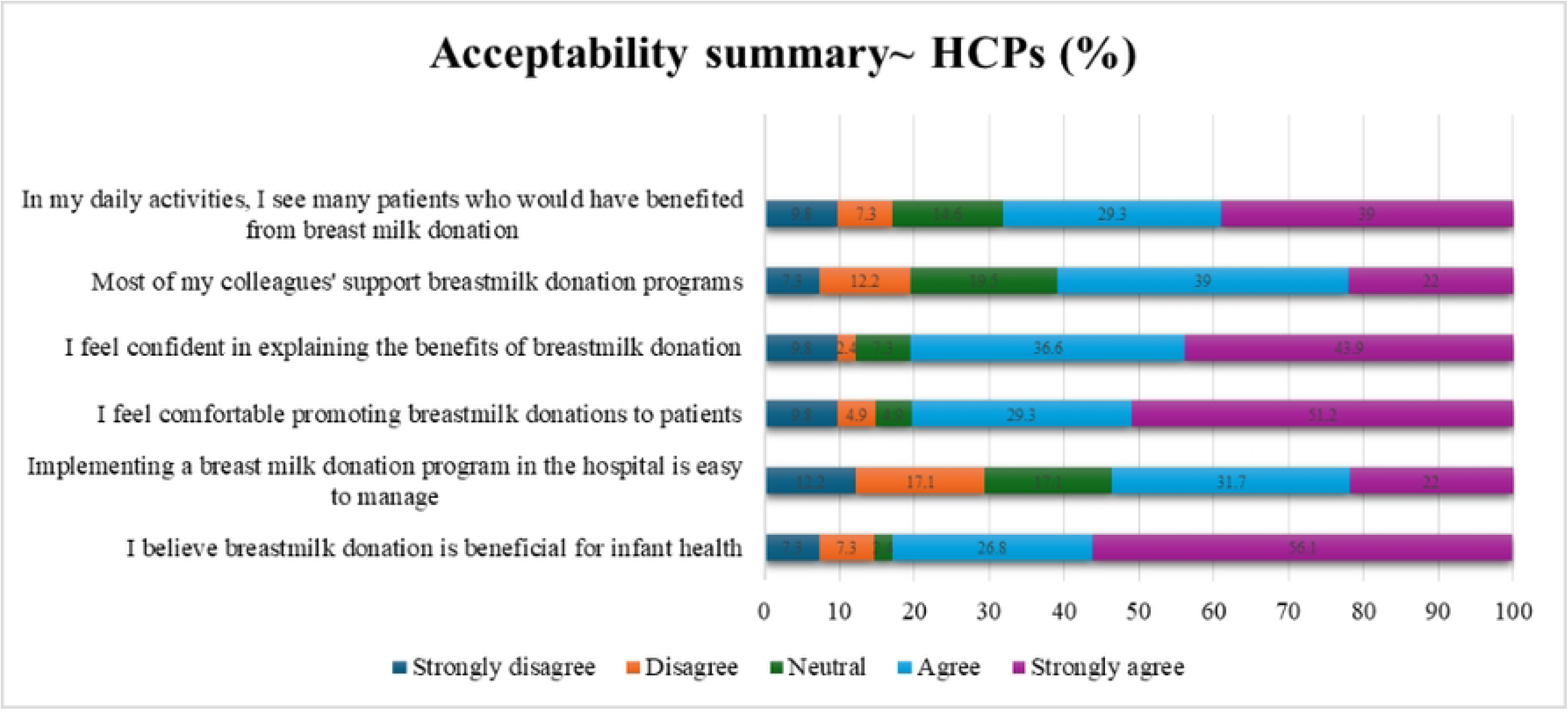
Summarizes the acceptability of breast milk donation among HCPs across six item

**Fig 2.**
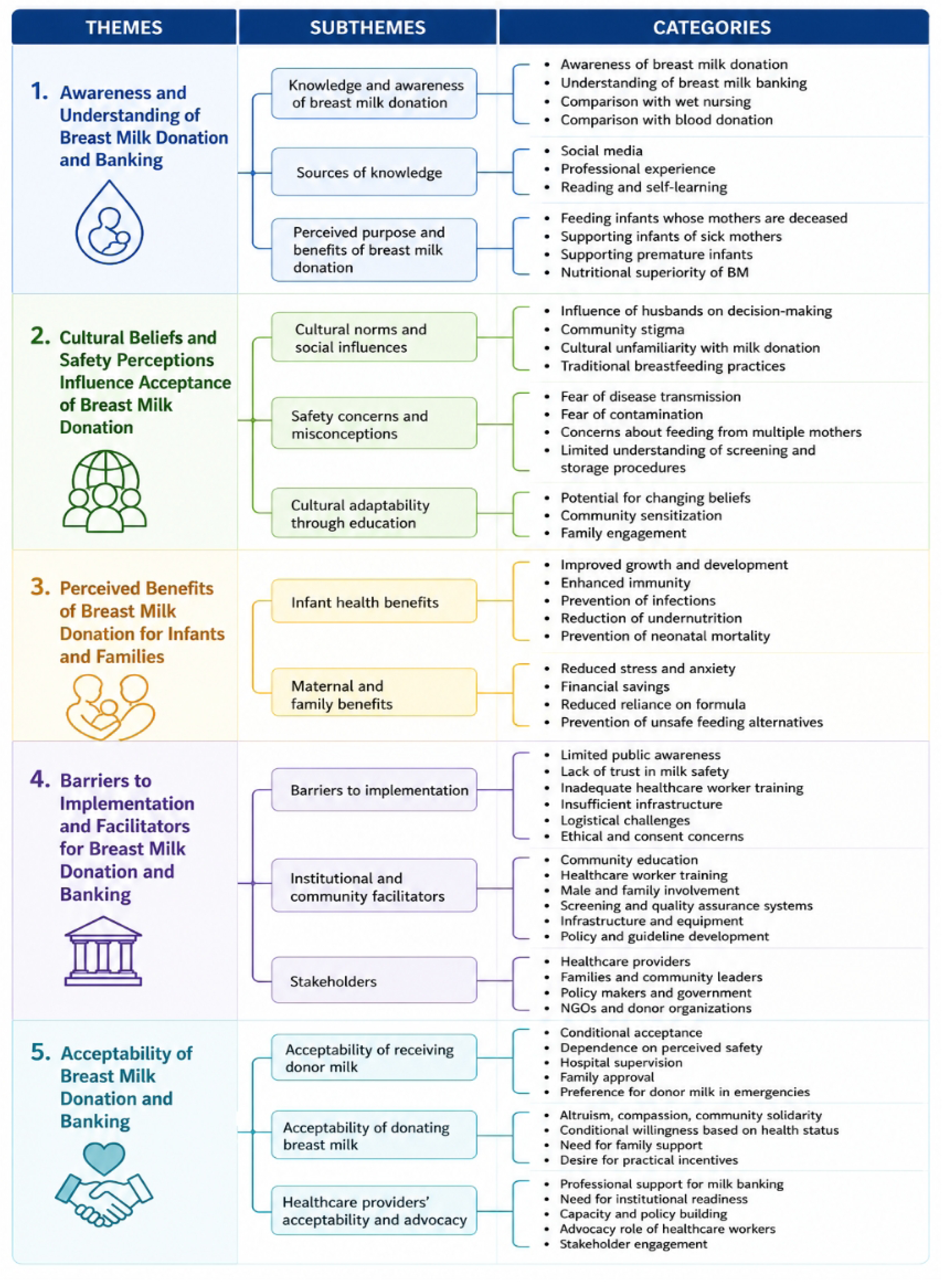
Summarize the themes, subthemes, categories, and codes that emerged from the analysis

Quantitative and qualitative findings were integrated during interpretation to identify converging evidence, with qualitative insights contextualizing quantitative patterns.

### Ethical Considerations and approval

This study was conducted in accordance with the ethical principles with references made to the Belmont Report and the Declaration of Helsinki. Ethical approval for the study was granted by the UGHE Institutional Review Board (Approval No. UGHE-IRB/2025/381), and permission was sought from the Ministry of Health, and the leadership of both hospitals.

The study did not pose any risk to participants. Participation in the study was voluntary, and written informed consent was obtained from all participants after full information about the study was provided, questions were answered, and willingness to participate was reconfirmed prior to signing *(S3 appendix).* Permission was sought before recording interviews, and written notes were taken when participants declined to audio recording. Confidentiality was ensured by collecting data anonymously and storing it securely, with no personal identifiers recorded. To protect confidentiality, only the participants and interviewers were present during each qualitative interview; no other staff, patients, or family members were in the room.

*S3 appendix: Written informed consent form used for study participants*

## RESULTS

### Quantitative results

#### Sociodemographic characteristics

A total of 41 healthcare HCPs surveyed, 20 (48.8%) were from KDH and 21 (51.2%) were from RL2TH. 3 (7.3%) were doctors, 31 (75.6%) were nurses, 4 (9.8%) were midwives, 2 (4.9%) were expert mothers, and 1 (2.4%) was a social worker. Most HCPs 27 (65.9%) had less than three years of professional experience in the neonatology department (Table 1).

**Table 1.** Healthcare Providers’ demographics.

| Variable | Categories | n =41 (%) |
| --- | --- | --- |
| Hospital | KDH | 20 (48.8%) |
|  | RL2TH | 21 (51.2%) |
| Job title | Doctor | 3 (7.3%) |
|  | Nurse | 31 (75.6%) |
|  | Midwife | 4 (9.8%) |
|  | Expert mothers | 2 (4.9%) |
|  | Social worker | 1 (2.4%) |
| Years of experience | <3 | 27 (65.9%) |
|  | >=3 | 14 (34.1%) |

#### Awareness and perspectives regarding breast milk donation among HCPs

Among HCPs 32 (78.0%) reported having prior exposure to the concept of breast milk donation and banking. 26 (63.4%) were somewhat familiar, while 10 (24.4%) indicated they were not familiar with the concept. Nearly all respondents, 40 (97.6%), believed that breast milk donation could be beneficial for neonates. Three-quarters, 31 (75.6%), agreed that there is cultural resistance to the donation of breast milk in the community. In terms of feasibility, 16 (39%) and 21 (51.2%) viewed the establishment of a breast milk bank as very feasible and feasible, respectively (Table 2).

**Table 2.**
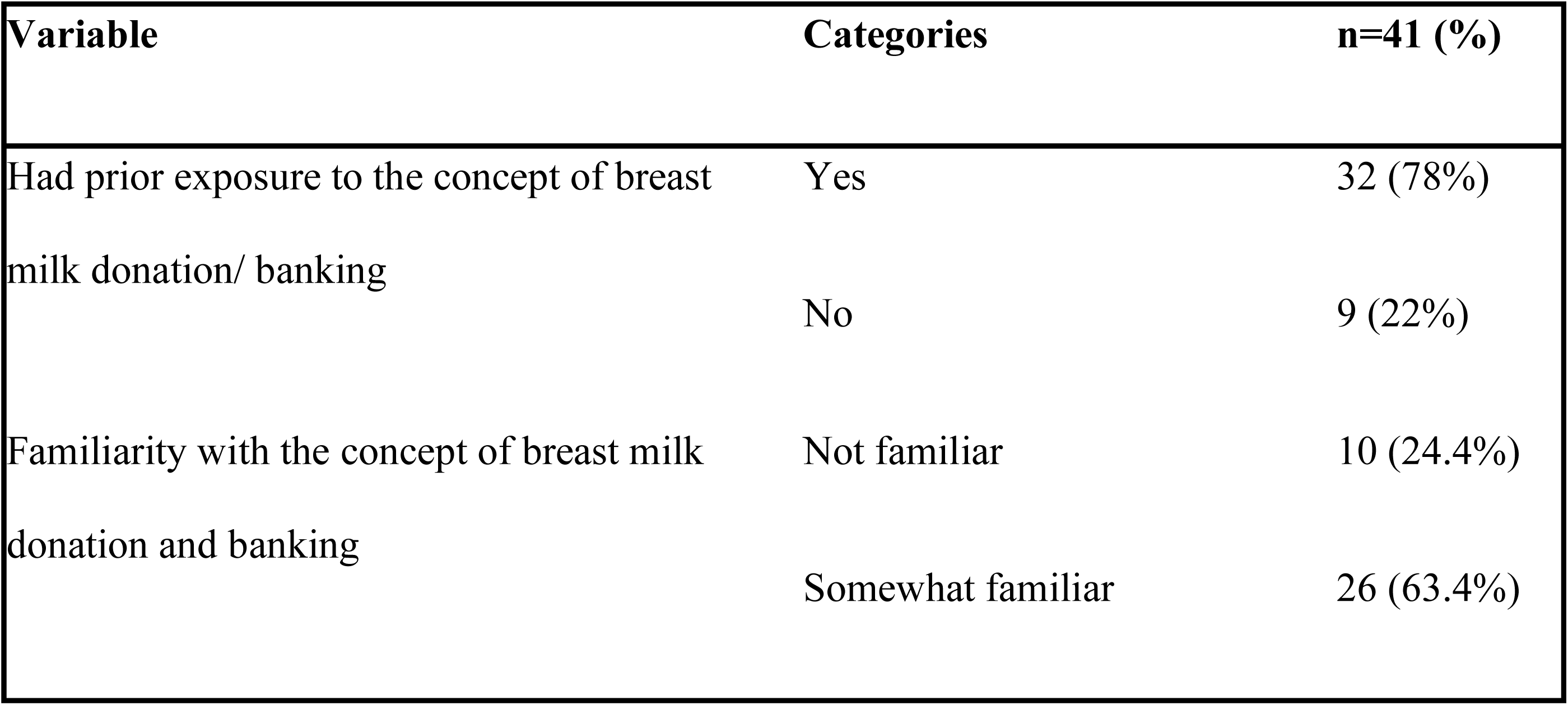
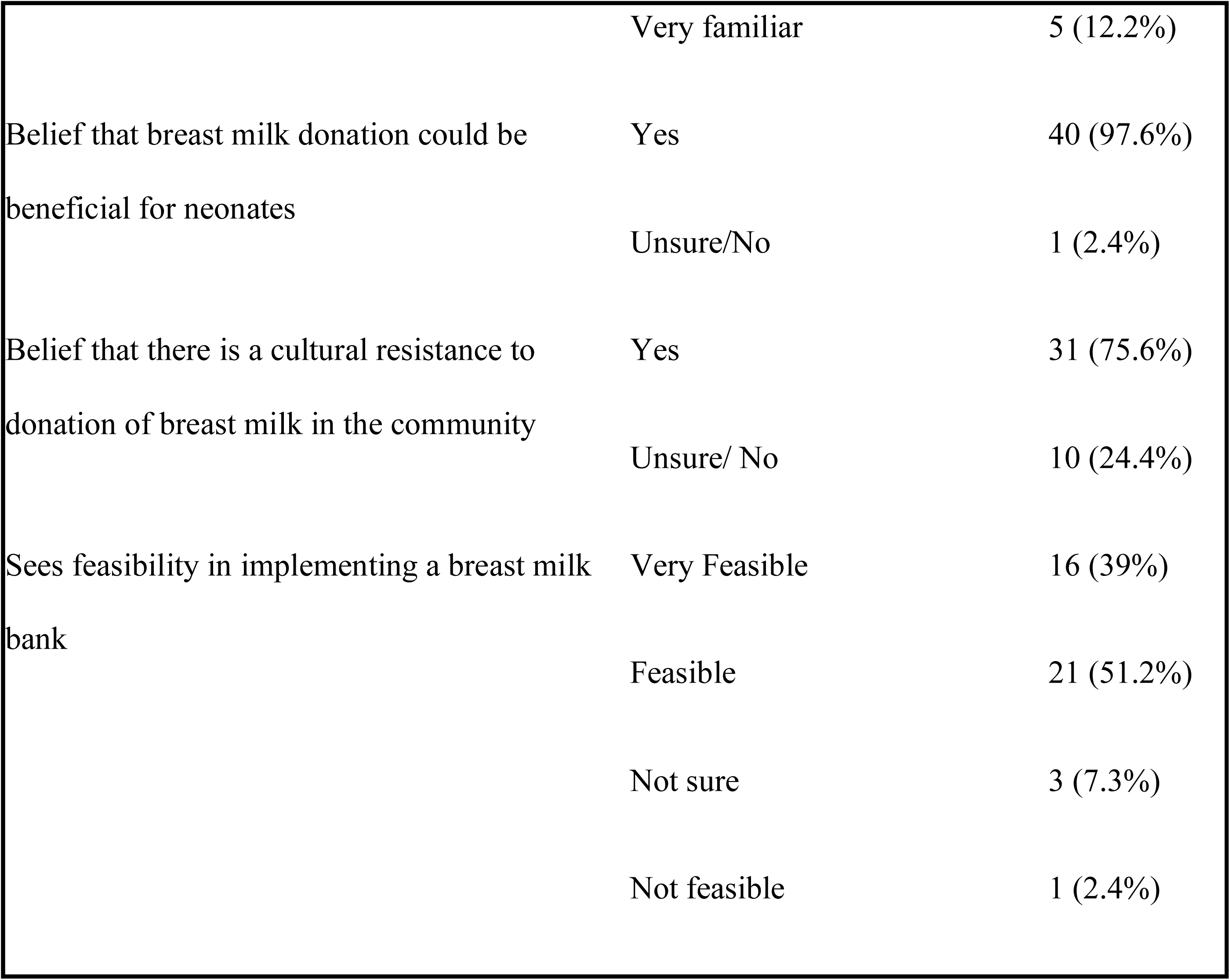
Awareness, perception regarding breast milk donation and banking∼ HCPs.

#### Acceptability to donate or receive breast milk

Overall, most participants expressed positive attitudes toward breast milk donation and banking practice. HCPs’ agreement with each statement ranged from 53.7% for seeing ease in managing the implementation of breastmilk donation program in their hospitals to 80.5% for feeling confident and comfortable promoting and explaining the benefits of breast milk donation program. The highest proportion of disagreement was observed for seeing ease in managing the implementation of breastmilk donation programs in their hospitals, with 29.3% (Figure 1).

Using a cutoff of 2.4, which marks the beginning of the “agree” range on the Likert scale, with acceptability scores dichotomized into “agree” (≥ 2.4) and “disagree/neutral” (< 2.4), the overall acceptability of the breast milk donation program among healthcare providers was high, with 82.9% (n = 34) reporting positive views. The mean acceptability score was 2.83 (SD = 1.10).

Comparisons of mean acceptability scores showed no significant differences across demographic or professional characteristics. There was no difference in acceptability between hospitals (p = 0.592), by years of professional experience (p = 0.751), or across job titles (p = 0.489). Overall, acceptability levels were consistent regardless of workplace, experience, or cadre (Table 3).

**Table 3.**
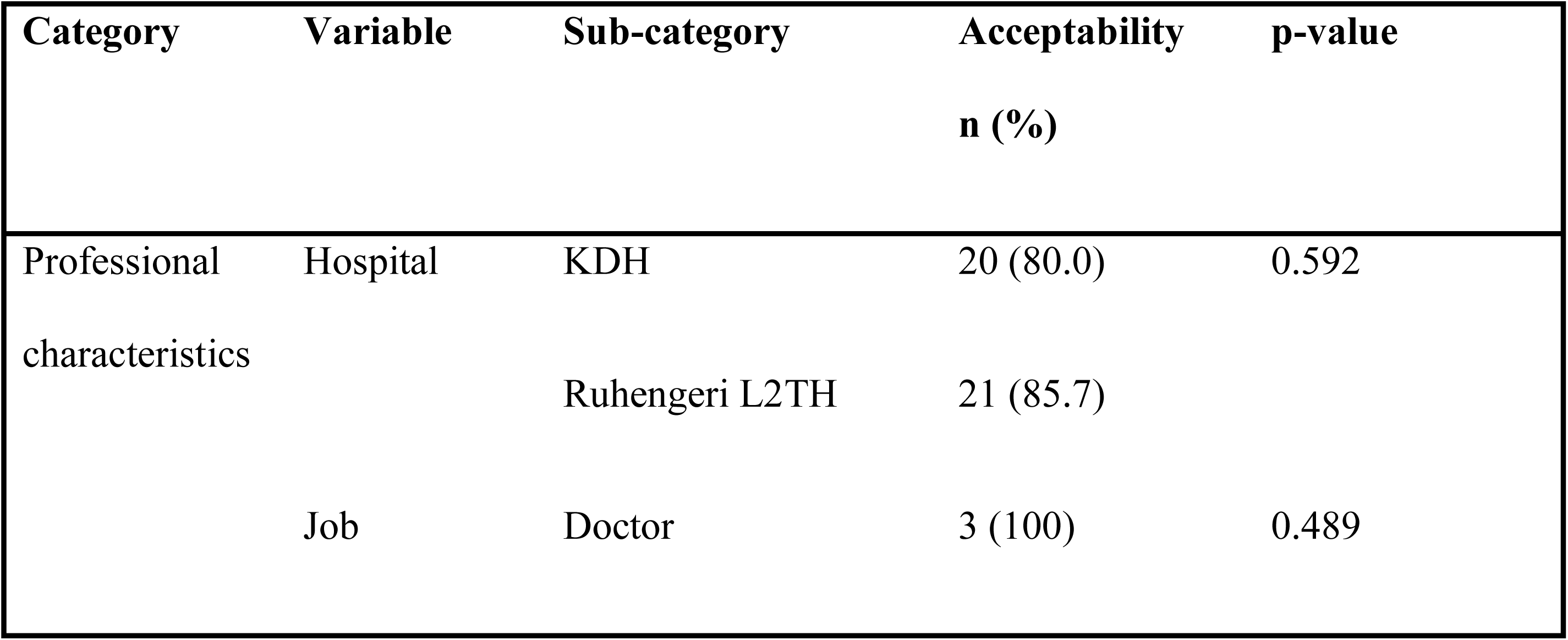
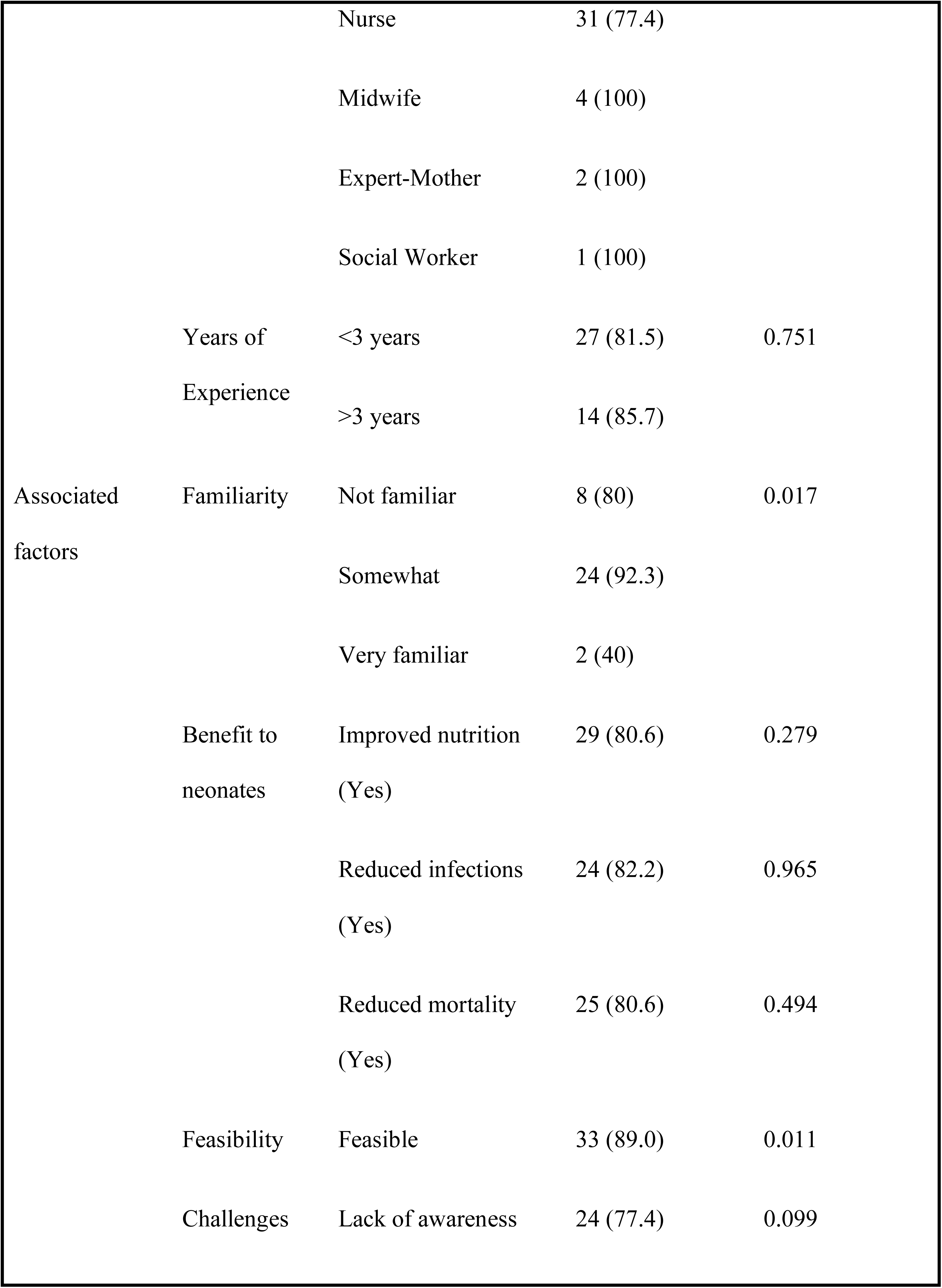
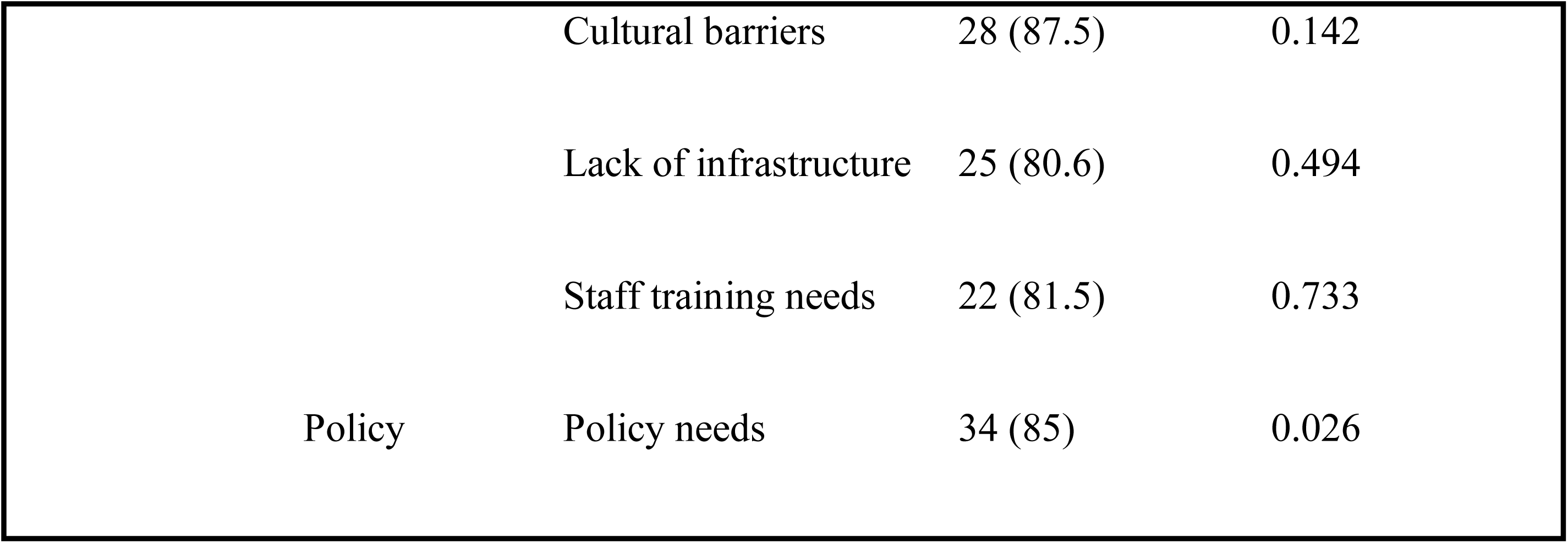
Determinants of Healthcare Providers’ Acceptability of Breast Milk Donation and Banking.

#### Factors Associated with Acceptability

Significant associations were found between acceptability and participants’ familiarity with breast milk donation (p = 0.017), feasibility perceptions (p = 0.011), and policy needs (p = 0.026). (Table 3).

#### Perceived benefits, barriers, and facilitators of breastmilk donation program

Healthcare providers highlighted several benefits of breast milk donation, including improved infant nutrition; 36 (87.8%), reduced neonatal mortality; 31 (75.6%), and reduced risk of infections; 29 (70.7%). Reported barriers included cultural concerns; 32 (80%), lack of awareness among mothers; 31 (77.5%), inadequate infrastructure for milk storage; 31 (77.5%), and staff training needs; 27 (67.5%).

As facilitators, all providers; 41 (100%), emphasized the need for training on how to promote and manage breast milk donation. They also highlighted institutional policies; 37 (90.2%), community engagement strategies; 37 (90.2%), and improved awareness; 36 (87.8%) as critical supports for successful implementation (Table 4).

**Table 4.**
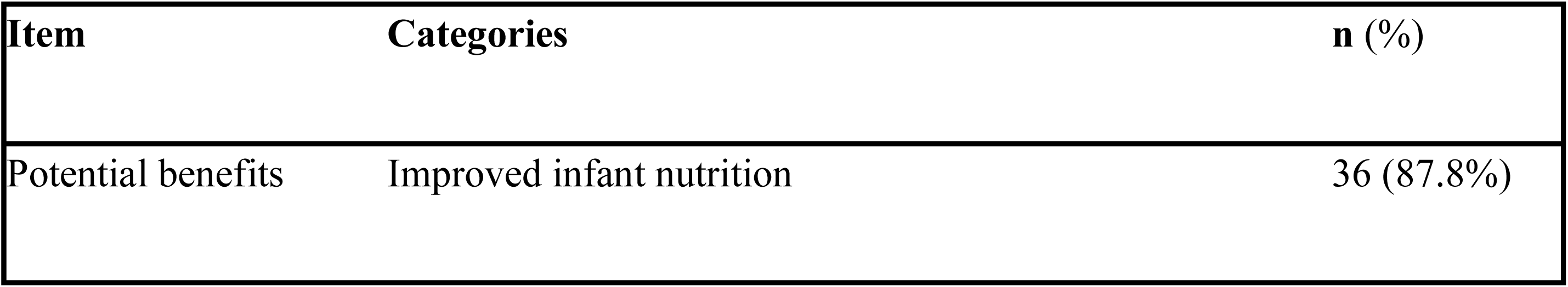
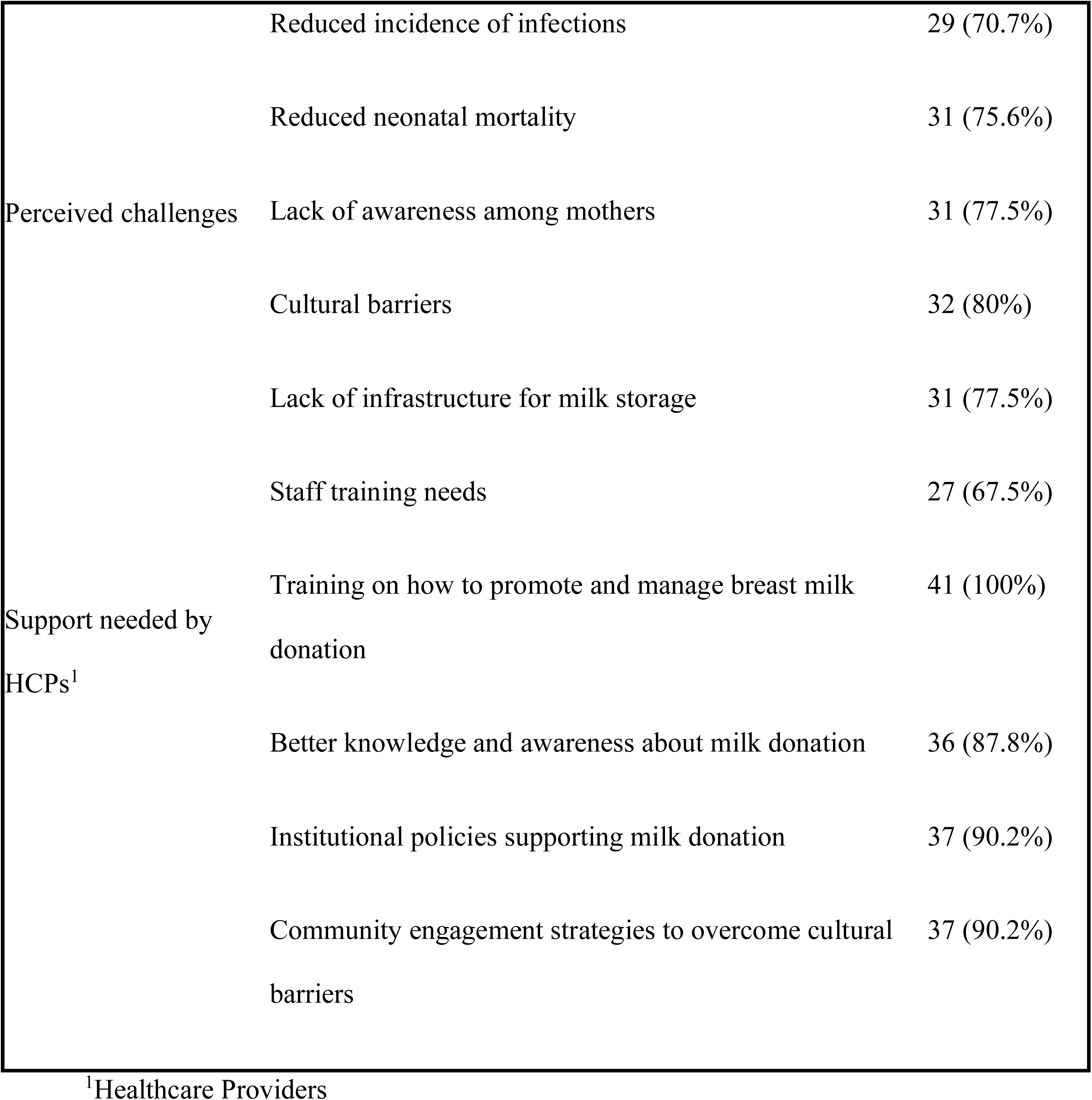
Perceived benefits, challenges, and facilitators

### Qualitative results

A total of 10 healthcare providers were interviewed (5 from each hospital): 1 doctor, 2 neonatal nurses, 1 expert mother, and 1 social worker at Kirehe District Hospital, and 1 doctor, 2 nurses, 1 midwife, and 1 social worker at Ruhengeri Level II Teaching Hospital; staff directly involved in neonatal care, irrespective of their years of experience. These interviews examined feasibility and operational considerations, institutional readiness, and anticipated challenges and enabling factors for implementing donor breast milk within the hospital settings (Figure 2).

#### Theme 1: Awareness and understanding of breast milk donations among healthcare providers

Some stated that they had learned about it through professional experience, reading, or hearing about similar programs in other countries. Healthcare providers mentioned that they knew the milk should be expressed, stored under hygienic conditions, and used for infants whose mothers are unable to breastfeed due to illness, death, or insufficient milk production. While awareness among healthcare providers was higher, both groups identified the absence of formal programs and limited national-level information as key barriers to wider understanding.

> Breastmilk donation and banking, I know that that exists in foreign countries, but to this day, there is no such program in Rwanda. ∼H2-03

> It can be helpful, sometimes the neonates are born but their mothers don’t have BM, or the mother is dead, or transferred to another hospital without the baby… therefore it can be helpful to help that child to have a healthy life. ∼ H1-01

> What I know about breast milk banking, we don’t have that in Rwanda. But I have read around. It is a banking, or service/department that prepares and stores breast milk. The breast milk is kept, designated to feed neonates that do not have their mothers, or whose mothers are sick. Such breast milk, as I read, helps such neonates, helping them to stay well nourished; as we know, breast milk contains natural ingredients to facilitate better growth. Secondly, since breast milk is well kept, it protects neonates from infections. ∼ H2-01

#### Theme 2: Cultural beliefs and misconceptions influence mothers’ views on breast milk donation and its safety

Healthcare providers acknowledge cultural and safety concerns. Many stated that these beliefs reflect limited information rather than strong opposition to milk donation. These accounts underscore that reluctance to breast milk donation stems possibly from limited understanding rather than cultural opposition, suggesting that targeted education could increase acceptance.

> As we know, Africans or even in Rwanda, breastfeeding is cultural. It is not practice, it does not normally exist, that someone feeds their neonate someone else’s breastmilk. But I think we can grow past such an understanding. Yes, our culture is beautiful, we can conserve it, but again, a culture that limits our development, we must avoid that… ∼H2- 01

> … but if there are teachings, the believes would change, because now people don’t understand it. It can be helpful. ∼∼H1-01

#### Theme 3: Healthcare providers recognized multiple benefits of breast milk donation for infant health and maternal emotional and financial assurance

Healthcare providers strongly endorsed the benefits of breast milk donation, describing it as highly advantageous for infants and mothers alike. For infants, participants emphasized that donated breast milk supports healthy growth, strengthens immunity, helps babies reach developmental milestones, and reduces the risk of infections, undernutrition, and neonatal death.

They also noted that donation can reduce financial strain for families who cannot afford formulas and prevent reliance on unsafe alternatives such as cow milk, offering both practical and emotional reassurance. Together, these findings indicate broad perceived benefits of breast milk donation, reinforcing its potential role in improving neonatal outcomes and supporting maternal wellbeing.

> *… for me I’m happy for this program because it can prevent mother from stress and anxiety and malnutrition for babies. And those formulars are expensive compared to the BM∼∼ H1-04*

> *Primarily because it would help the parents who opt for infant formulas. Breastmilk contains nutrients and antibodies that can’t be found in infant formulas. ∼H2-04*

*Theme 4: Successful Implementation of Breast Milk Donation Programs Depends on Awareness, Trust, and Institutional Support*

### Subtheme 4a: Limited Awareness, Infrastructure, and Trust Reduce Program Feasibility

Healthcare providers shared concerns, emphasizing gaps in public knowledge and limited professional training around breast milk banking. They noted that without clear institutional guidelines, proper storage facilities, or established laboratory procedures for screening and pasteurization, implementation would remain challenging.

> *Barriers are there like infrastructures, that bank, also teaching the mothers that will donate, and also healthcare providers having no knowledge about it can be a barrier ∼H1-03*

> *The primary challenge is material hygiene, materials to express in, hygiene of where the milk will be kept, all of that, it requires a lot of material resources, fridges to keep them, and then other materials to warm them before giving them to the baby. ∼H2-04*

> *Even ourselves, the hospital workers, there is a lot we do not know about the program. It would require training for us to also understand. ∼H2-05*

#### Subtheme 4b: Education, institutional support, and community engagement can facilitate the safe and acceptable implementation of breast milk donation programs

Despite these barriers, healthcare providers identified practical strategies that could facilitate the successful introduction of breast milk donation in Rwanda.

Healthcare providers highlighted the need for institutional support through professional training. They also recommended materials for proper handling and storage systems.

> *I think it’s okay, but also make sure that the BM, which is kept, is kept in a safe place that they won’t cause any harm to the baby, …” ∼ H1-02*

> *The primary support is about hospital staff, then training the hospital staff, followed by training mothers as well as the community for them to understand that such a program exists. Then the hospital would need materials that will be used to prepare the breast milk donated. ∼H2-03*

Healthcare providers agreed that collaboration and following hospital-level policies/guidelines to implement the program would increase public confidence. A few also suggested providing nutritional or logistical support for donor mothers to encourage participation.

> *I think we could all work together to train the community, but only after have we also been trained. Another contribution would be to adhere to the established programs, following the guidelines that would rule the donation and storage of breastmilk and give it as we have been trained. ∼H2-02*

While most themes reflected broad convergence, the degree of confidence in feasibility varied: as captured in Subtheme 4a, some providers voiced more cautious views, citing infrastructure and training gaps (e.g., ∼H1-03, ∼H2-04), while others expressed stronger optimism about implementation.

#### *Theme 5: Healthcare Providers Link Acceptability to Institutional Capacity and Program* Feasibility

Healthcare providers supported breast milk donation but emphasized that their willingness to participate and promote it depended on institutional capacity. They stressed that all staff should be trained to support mothers directly. Also, providers noted that acceptability would be strengthened when staff are confident in existing systems, including storage and testing, and when hospitals foster ongoing education and engagement with both staff and stakeholders.

> *First is that every HCWs should have the knowledge, so that they can support the mothers they met, without waiting for specific people to come and do the teaching. ∼ H1- 02. In hospital we need many things, advocacy, because I think we will need other materials so that it can happen, and people also need a lot of teaching and also, we will need stakeholders ∼ H1-05*

These findings suggest that to establish a breast-milk bank in Rwanda (or similar settings), attention must be given to awareness-raising, culturally sensitive education, trust in screening/safety, and the role of healthcare providers as advocates.

> *In hospital we need many things, advocacy, because I think we will need other materials so that it can happen, and people also need a lot of teachings and, we will need stakeholders ∼ H1-05*

## DISCUSSION

The study specifically explored the acceptability of breast milk donation for neonatal feeding among healthcare providers in two Rwandan district hospitals. Overall, participants showed high levels of acceptability, reflecting broad recognition of donor milk’s potential to improve neonatal health outcomes. These findings are consistent with studies in other low- and middle-income countries showing positive attitudes and growing support for donor milk initiatives when communities are adequately informed [35], [36].

Quantitative findings demonstrated generally high acceptability of breast milk donation among healthcare providers, with strong agreement that donor milk benefits vulnerable infants and is safe when its handling, screening, and pasteurization procedures are clearly explained. Acceptability did not vary significantly across most demographic characteristics. However, the results also revealed important gaps; limited awareness and knowledge, safety concerns, fear of community resistance, and insufficient health system preparedness.

Healthcare providers expressed strong motivations that could facilitate adoption: a shared belief in the health benefits of breast milk for vulnerable infants, trust in biomedical processes, perceived financial relief compared to formula, and cultural values of helping others. Concerns were largely centered on safety, hidden infections, and limited health-system readiness, rather than intrinsic opposition to the idea of donating or receiving milk.

Together, these quantitative and qualitative findings demonstrate that donor milk is broadly acceptable in Rwanda when concerns about safety, awareness, and institutional capacity are addressed. This alignment across data sources is illustrated in Figure 3, highlights how both strands converge around similar facilitators and barriers.

**Fig 3.**
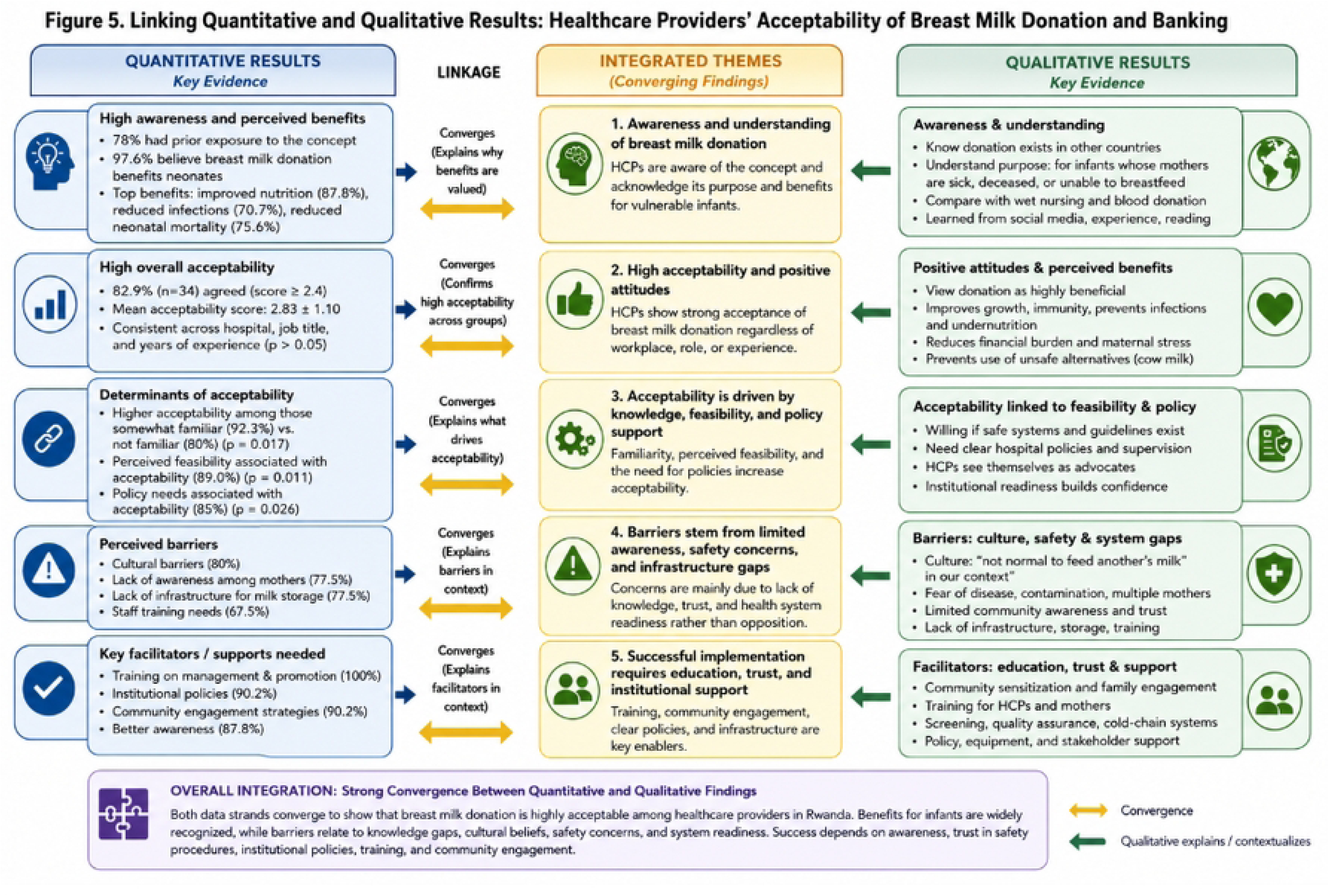
Link between quantitative and qualitative themes

HCPs demonstrated greater conceptual understanding and familiarity but highlighted the absence of structured system or training to guide milk donation or storage, echoing constraints reported in other African settings where infrastructure and policy gaps hinder adoption and implementation [15], [18].

Importantly healthcare providers emphasized that beliefs stem primarily from lack of knowledge rather than outright rejection of milk donation, underscoring the need of culturally sensitive education. Participants strongly agreed that donor milk improves infant nutrition, reduces infections, and lowers neonatal mortality. These perceptions are in line with global evidence regarding donor milk’s protective effects against NEC and other morbidities, especially for preterm and low-birth-weight infants [37]. Additionally, HCPs described emotional and financial relief as secondary benefits, including reduced stress related to feeding difficulties and the high cost of formula. These broader psychosocial and economic dimensions align with global findings that donor milk contributes to maternal well-being and house stability, especially in low resource settings [38], [39].

Barriers identified in this study; low awareness, cultural stigma, safety concerns, and limited infrastructure; aligned with evidence from other sub-Saharan African contexts [35], [36]. HCPs emphasized gaps in cold-chain facilities and standard operating procedures as key obstacles to implementation. Nevertheless, participants agreed that awareness, professional health support, and clear institutional policies could substantially enhance acceptability. This mirrors lessons from Brazil’s successful milk bank network, where strong policy backing and systematic training contributed to successful implementation [40], [41], [42], [43].

Quantitative analysis revealed HCPs, familiarity with the concept and perceived institutional feasibility shaped acceptability, highlighting the need for capacity-building and policy support. HCPs expressed high willingness to support milk donation but emphasized that their participation would depend on institutional readiness, availability of guidelines, and training.

Overall, these findings suggest that establishing a breast milk bank in Rwanda, and similar settings, requires coordinated efforts across awareness-raising, culturally sensitive community engagement, assurance of safety and screening, and strengthening of institutional capacity. HCPs are positioned to play a critical role as advocates, but their effectiveness will depend on clear policies, training, and infrastructure. The convergence of motivations and barriers among providers highlights a strong foundation upon which national and facility-level systems can be built to promote donor breast milk for neonatal care.

### Limitations of the Study

While this study provides important insights into the acceptability and determinants of breast milk donation and banking among healthcare providers in two Rwandan hospitals, several limitations should be acknowledged. First, the study was conducted in only two district hospitals, which may limit the generalizability of the findings. Although these hospitals serve diverse rural populations and represent typical Rwandan neonatal care contexts, perspectives from urban or private hospitals may differ. Second, participants had no direct experience with breast milk donation and relied on theoretical knowledge for their responses; their views may therefore differ from those formed through actual practice or exposure to milk banking programs.

Furthermore, the cross-sectional design and the absence of existing milk banks limit the ability to understand how acceptability might evolve over time. Participants’ views were based on a hypothetical scenario, and their perceptions may change with real-world exposure to education, counselling, or actual implementation of donor milk banking. Additionally, transcripts and findings were not returned to participants for member-checking, which may have offered further validation of the interpretations presented.

Finally, while this study explored acceptability and perceptions of breast milk donation and banking, it did not include a comprehensive assessment of logistical feasibility, which warrants further investigation through dedicated implementation or health systems research.

Despite these limitations, this mixed methods study provides foundational evidence to inform policy, planning, and early implementation of breast milk donation and banking programs in Rwanda. A key strength of this work is its methodological approach: because this is a novel area of research, a mixed methods design was necessary to capture quantitative trends as well as the qualitative reasons behind them, insights that would not have been possible using only a single quantitative or qualitative approach.

## CONCLUSION

Despite limited awareness and reliance on theoretical knowledge rather than direct experience, providers expressed high acceptability, recognized clinical benefits, and indicated willingness to participate in donation programs. Cultural beliefs, perceived safety, and understanding of the health benefits all play a significant role in shaping attitudes toward donor breast milk. This study demonstrates that HCPs generally perceive breast milk donation as a valuable intervention to improve neonatal nutrition, reduce morbidity, and support vulnerable infants. Key determinants of acceptability included perceived benefits for infant survival, positive attitudes toward milk sharing, belief in the feasibility of establishing milk banks, and willingness to participate. However, concerns such as cultural norms, lack of knowledge, and inadequate hospital infrastructure remain important barriers. The findings contribute essential baseline information for policy formulation, preparatory planning, and advocacy for integrating donor breast milk into Rwanda’s neonatal health strategy. Importantly, before any pilot milk bank is established, a comprehensive needs assessment will be required to determine the extent of need and guide appropriate resource allocation, ensuring that implementation responds to actual demand.

## Data Availability

Due to the sensitive nature of the data and the small sample sizes at each study site, the minimal dataset underlying this study's findings cannot be made fully publicly available without risking participant re-identification, which was not authorized under the informed consent obtained or the approval granted by the UGHE Institutional Review Board (Approval No. UGHE-IRB/2025/381). De-identified data may be made available to qualified researchers upon reasonable request and with permission from the UGHE Institutional Review Board. Requests can be directed to the UGHE IRB or the corresponding author.

## LIST OF ABBREVIATIONS

BM: Breast Milk
HCP(s): Healthcare Provider(s)
KDH: Kirehe District Hospital
NEC: Necrotizing Enterocolitis
RL2TH: Ruhengeri Level II Teaching Hospital
SPSS: Statistical Package for Social Sciences
UGHE: University of Global Health Equity
UNICEF: United Nations Children’s Fund
WHO: World Health Organization

## ACKNOWLEDGEMENTS

We extend gratitude to our supervisors, Dr. Ibrahim Olayinka and Dr. Augustine Ndaimani, for their guidance, mentorship, insights, and support. We are equally grateful to our preceptor, Dr. Peace Kakibibi, for her consistent encouragement and insightful feedback.

Our appreciation goes to Prof. Rex Wong and Prof. Chester Kalinda, whose expert input in data analysis and constructive feedback helped refine this report.

We thank our data collectors, Mr. Zachee Niyomugabo and Ms. Solentine Umunezero, for their commitment and diligence.

We are grateful to the administration and staff of Ruhengeri Level II Teaching Hospital and Kirehe District Hospital for their collaboration and support.

Special thanks to Dr. Augustin Tuyishime, whose insightful discussions on issues surrounding breastmilk sharing inspired us to pursue this research topic.

Finally, we thank the University of Global Health Equity for transformative education, mentorship, opportunities, and deep sense of social responsibility that made us passionate global health leaders.

